# Histopathology: Deep machine learning based semantic segmentation features predict patient survival

**DOI:** 10.1101/2023.01.14.23284554

**Authors:** Vikas Ramachandra

## Abstract

In this paper, we use deep learning techniques to segment different regions from breast cancer histopathology images, such as tumor nucleus, epithelium and stromal areas. Then, in the second stage, the deep segmentation features learned by the neural network are used to predict individual patient survival, using random forest based classification. We show that the deep segmentation network features can predict survival very well, and outperform classical computer vision based shape, texture and other feature descriptors used in earlier research for the same survival prediction task.

## 1. Introduction

Breast cancer has recently been classified as one of the top 5 fatal types of cancer in the world by the World Health Organization. In 2018, nearly 330,080 new cases of breast cancer (invasive and non-invasive) and 40,920 deaths were reported in the U.S [3]. In order to assess the stage and the severity of breast cancers, histological examination of tissue specimens, obtained through biopsy, remains the gold standard for diagnosis and accurate evaluation of breast diseases. Generally, the pathologists often use the Nottingham Histology Score (NHS), also referred to as the Scarff-Bloom-Richardson grading system [2], to classify and recommend the treatment strategy for breast cancers. Morphological features of breast cancer epithelial cells, such as tubule formation, epithelial nuclear atypia, and epithelial mitotic activity are each scored qualitatively (manual/visual inspection by the pathologist), and the assessments are combined to stratify breast cancer patients into different groups that have shown significant survival differences historically. Addition of new tests and biomarkers have improved this semi-quantitative morphological scoring scheme over the years but still remains the standard technique for histologic grading in invasive breast cancer. To further confirm the diagnosis, the image inspection based features can be augmented with sequencing and molecular data. But most diagnostic labs around the world still use pathology images as the first step in diagnosis.

The current manual image based grading system has a few limitations, wherein the concurrence is low and the variability in histologic grading among pathologists is high, which may lead to potentially negative consequences for treatment decisions. The introduction of digital pathology helped in dealing with huge and complex amount of information obtained from tissue specimens. Combined with whole-slide-imaging, computational digital pathology has proved to be very useful in accurate, precise and efficient image analysis of these tissues. Automated image analysis and computer vision algorithms help mitigate the subjectivity between the pathologists and provide an objective method for predicting patient prognosis directly from image data. Given the rapid evolution of these computer vision algorithms, computational digital pathology can accurately measure breast cancer morphologic characteristics, allowing objective stratification of breast cancer patients on the basis of morphologic criteria and facilitate the discovery of novel morphologic and prognostic features associated with response to specific therapeutic agents.

In the past, research has focused on using hand crafted computer vision based feature extraction from these images, and using machine learning to predict survival using the hand crafted feature set [1]. Recently, deep learning based techniques for image classification and segmentation have become popular for natural image datasets. In this paper, we use one such popular technique, and show that it works well for pathology image segmentation. Further, we extract features from the trained deep model, and build a classifier which shows good predictive accuracy for survival, and performs better than computer vision based hand crafted features which have been used in earlier papers for predicting survival, on this dataset. Thus, this paper shows that the use of deep learning based semantic segmentation and classification is a useful tool for estimating survival using pathology images alone.

## 2. Details of the dataset

Data was obtained from the Stanford Tissue Microarray Database (TMAD) and consists of microscopic images from two independent cohorts of breast cancer patients [from the Netherlands Cancer Institute (NKI) cohort, n = 248, and the Vancouver General Hospital (VGH) cohort, n = 328]. Manually drawn annotations/ masks were provided for each patient’s image, which contains region level details. Tumor, stroma, peri-tumor regions were marked with different colors on the mask, which is used as ground truth to train the segmentation algorithm. The data also consists of patient level survival scores.

## 3. Deep learning based segmentation

The first step of our algorithm is to train a semantic segmentation neural network, which can identify and segment different regions in the histopathology image, specifically: stroma, nucleus, background and epithelium regions. For this, we train the following network, whose architecture is shown in the figure below. The network consists of a convolution layer, followed by ReLu, max pooling, and second convolution and ReLu, and then upsampling, followed by a softmax+pixel classification layer which learns the output segmentation mask.

**Figure:**
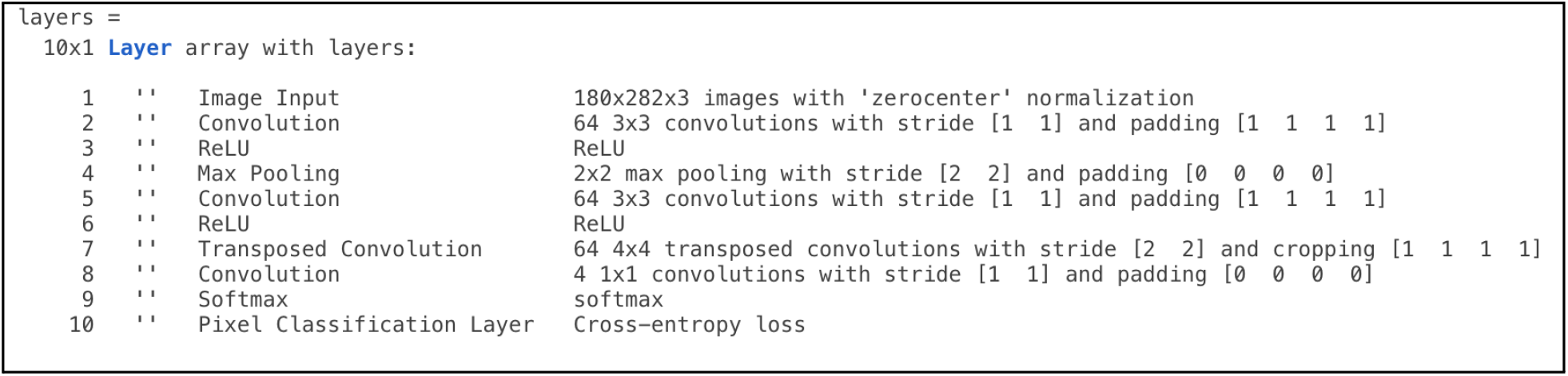
Network architecture

For training, all images are resized to 180×282×3 for faster training, and cross entropy loss is used as the metric. SGDM is used, with a learning rate of 0.001 and 100 epochs are run, with a minibatch size of 64 images. Since the classes are imbalanced, case weights are proportional to the relative class frequencies, as follows, for [background, stroma, epithelium, nucleus] : [1.045, 80.2759, 40.0267, 302].

The training losses and accuracies are as shown below, for the semantic segmentation network.

**Figure:**
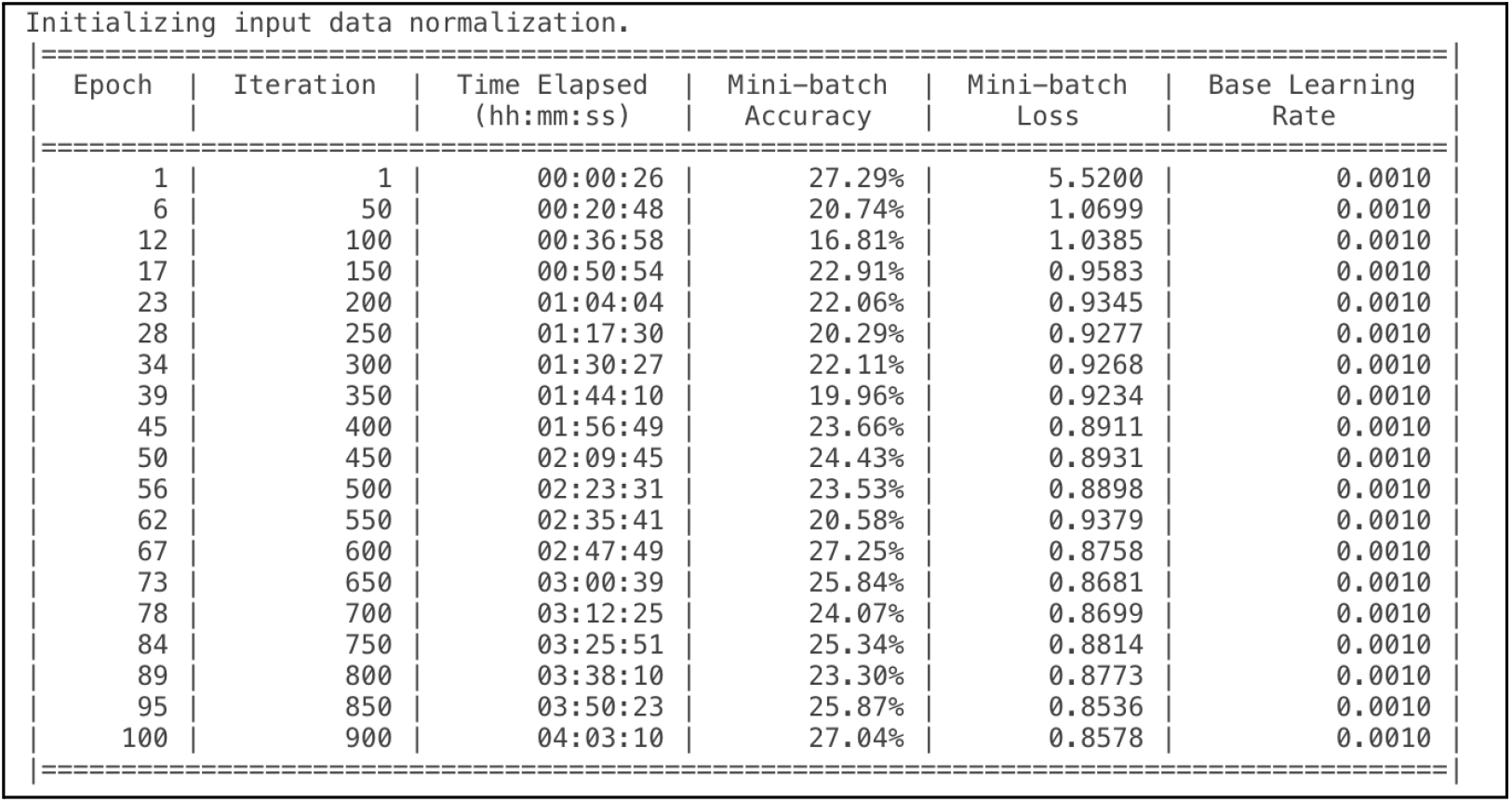
Training metrics for the semantic segmentation neural network (Segnet)

The above network is trained on the first set of patients (NKI patient cohort), and tested on the second cohort (VGH patient cohort).

An example of an input pathology image and the segmentation mask learned is shown in the figure below.

**Figure:**
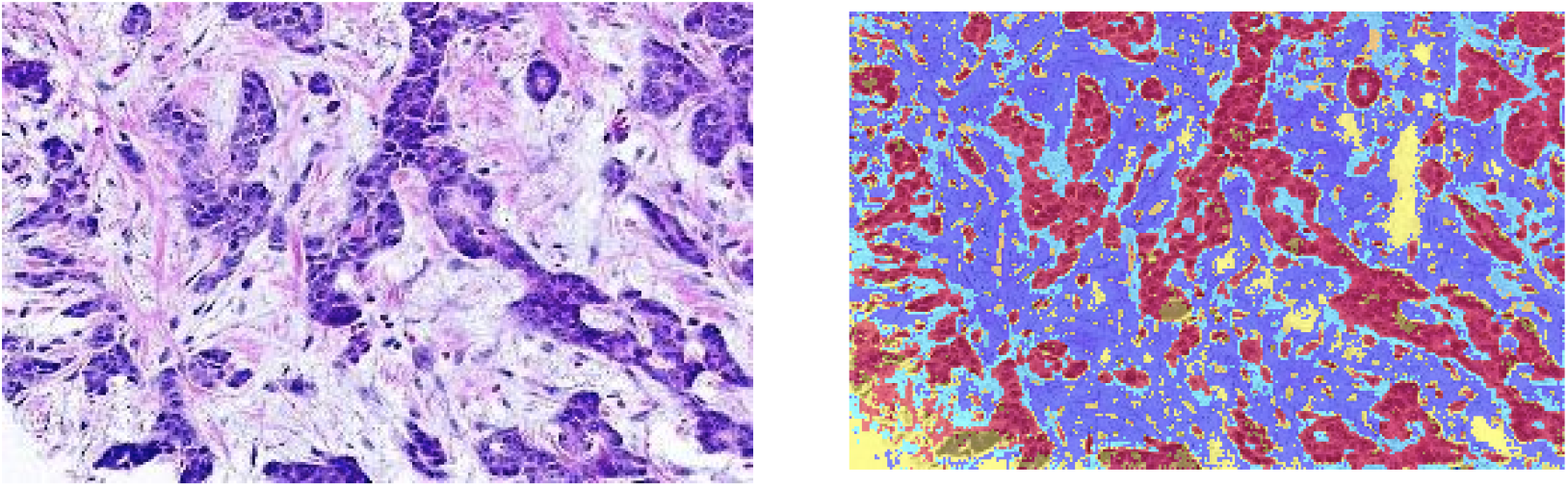
Input image: left and learned segmentation mask: right (different colors indicate different types of tissue)

Once we have the trained semantic segmentation network, we run the network in inference mode on the NKI patient cohort, and extract features as follows: the final 2 layers from the network (softmax and pixel classification layers) are removed, and the output from the network at the third convolution layer is extracted, This can be thought of as a way to featurize each image by projecting into the learnt latent space.

## 4. Survival prediction using segmentation features

Form the above step, the latent space features were obtained for each image. This feature set is then passed to a random forest classifier. The outcome ground truth labels for the classifier are patient survival (binary 0/1 whether the patient survived or not, for the follow up period). The random forest was trained on features extracted from the NKI cohort (on which the semantic segmentation network was trained), and 10 fold cross validation was used to avoid overfitting. For testing, we used the VGH cohort images. First, the latent space features were extracted for this cohort by running the neural network in inference mode (without the last 2 layers, as described above), then the features were passed to the trained random forest, which can classify the survival outcome based on the segmentation network features. The ROC curve and confusion matrix for this segnet features +Random forest are shown below.

**Figure:**
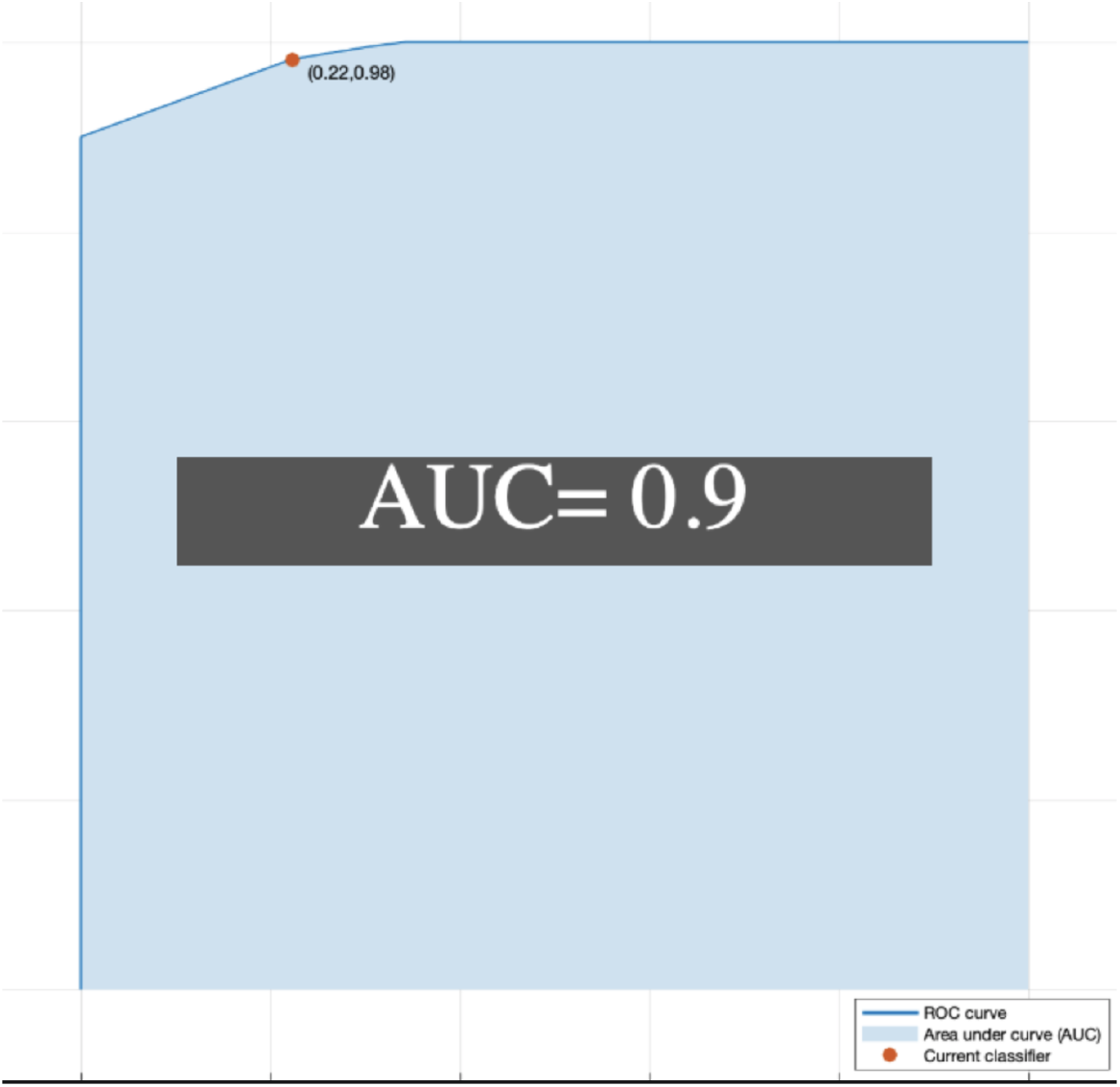
ROC curve for the segnet features plus random forest classifier for survival (Y axis: True positive rate, X axis: False positive rate. For an example operating point, TPR=0.94 with FPR =0.2)

**Figure:**
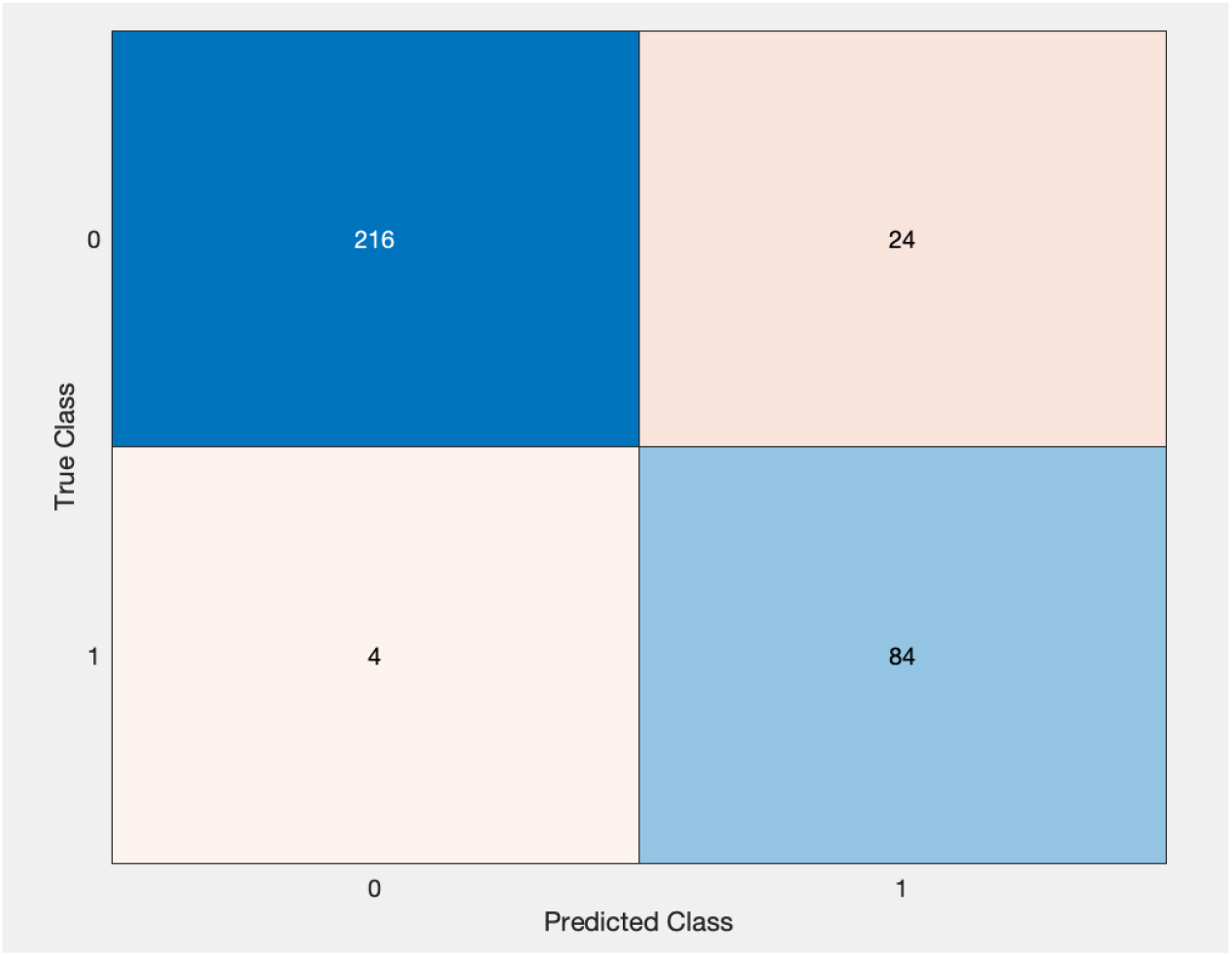
Confusion matrix (for the two classes, patient survived=0, died=1)

**Figure:**
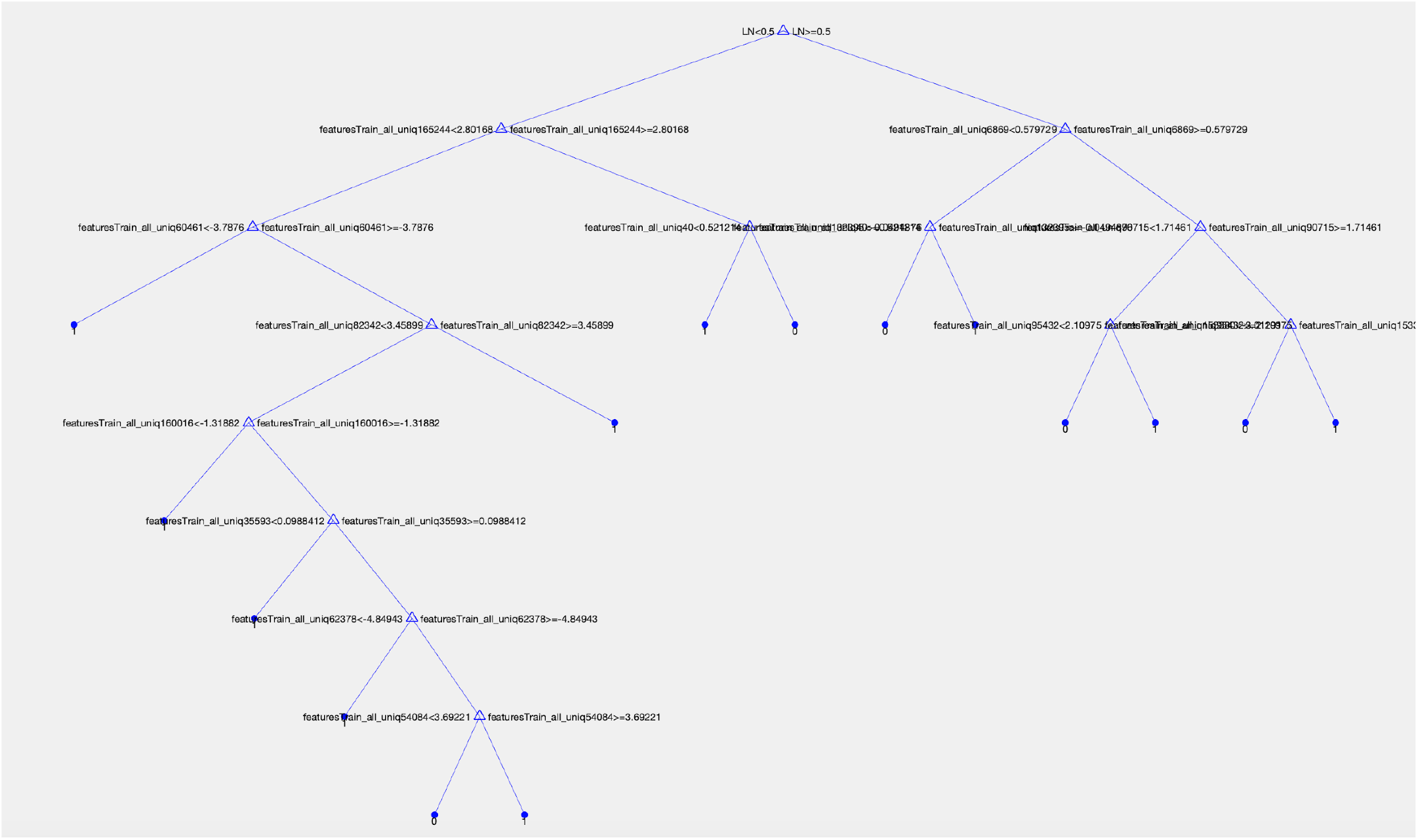
Example decision tree for the survival classifier (note that several of the nodes are features from the semantic segmentation network)

## 5. Discussion and conclusion

In this paper, we have developed a deep learning based algorithm that can predict the survival of breast cancer patients, from the features extracted from semantic segmentation of H&E pathology images. Our approach has successfully done semantic segmentation and extracted features from different tissue classes and correlate these tissue patterns to the survival outcomes. We have achieved a high AUC of 0.90. Our study provides insight into using novel deep learning algorithms to stratify and predict patient survival using only histopathology H&E images, bypassing other molecular and genetic testing that may be needed. Although our method has performed well on curated Tissue Microarray (TMA) data, its performance on large WSIs should be tested and evaluated in further studies. Also, the performance of our model should be evaluated and refined on larger patient populations from diverse cohorts, which will improve the predictions and generalizability of the model. The goal of this study is to demonstrate a proof of concept system to predict survival only from imaging data.

## Data Availability

All data produced are available online

## References

[1] Systematic Analysis of Breast Cancer Morphology Uncovers Stromal Features Associated with Survival; A.H. Beck et. al, 2011.

[2] Histological grading and prognosis of breast cancer.; H.J.G. Bloom, and W. W. Richardson, 1957. Br. J. Cancer 11, 359–377. doi: 10.1038/bjc.1957.43

[3] Breastcancer.org U.S., 2018

